# Impact of transportation on the suitability of corneal lenticule for implantation

**DOI:** 10.1101/2024.10.21.24315848

**Authors:** Jenetta Y. W. Soo, Gabriel Tan, Evelina Han, Kenny P. Y. Boey, Yu-Chi Liu, Jodhbir S. Mehta, Andri K. Riau

**Author notes:** Corresponding authors: Prof. Jodhbir S. Mehta Dr. Andri K. Riau.

## Abstract

**Background:** Corneal lenticules can be banked and retrieved for vision-restoring surgeries. Extended transportation logistic delays from the lenticule bank to the clinic could be a concern. This study investigated the effects of transportation on the lenticules.

**Methods:** Lenticules were cryopreserved at a Ministry of Health-licensed lenticule bank for 1 year and were transported at 4°C. The transparency was measured daily until significant degradation was notable, compared to fresh lenticules from donor corneas (n=3). The molecular and ultrastructural integrity of lenticules after 1 day in transport (n=3) and on the day of transparency deterioration (n=3) was evaluated by histochemistry and transmission electron microscopy (TEM). In addition, 6 rabbits were implanted with these lenticules to assess the difference in postoperative outcomes by corneal imaging, immunofluorescence staining, and TEM.

**Results:** Lenticules showed a significant transparency reduction after 6 days in transit (p=0.002). The change was not caused by molecular alterations but by a greater distribution shift in the interfibrillar distance (IFD) (Z=4.419; p<0.001) and fibrillar diameter (FD) (Z=6.435; p<0.001). Post-implantation, day 6 lenticules exhibited greater haze and slower recovery of clarity compared to fresher lenticules, despite corneal imaging and immunofluorescence staining showing no fibrosis, inflammation, or vascularization in either group. With TEM, the discrepancy was revealed due to the difference in the recovery of IFD and FD distribution.

**Conclusions:** Banked lenticules can maintain transparency for up to 5 days of transportation at 4°C. Further delays compromise their ultrastructural integrity and postoperative clarity, emphasizing the need to factor in transportation in lenticule banking logistics.

## Introduction

Refractive surgery, which includes Laser-Assisted In Situ Keratomileusis (LASIK), Photorefractive Keratectomy (PRK), and Keratorefractive Lenticule Extraction (KLEx), are popular methods to correct refractive errors worldwide, especially in Asia due to its high rates of myopia (1). One of the earliest KLEx procedures, Small Incision Lenticule Extraction (SMILE; Carl Zeiss Meditec, Jena, Germany), has been a popular method of surgery ever since its introduction in 2010 due to its minimally invasive nature, and reduced risk of flap-related complications (2). In the past 3 years, more KLEx procedures were introduced, namely Corneal Lenticule Extraction for Advanced Refractive correction (CLEAR; Ziemer, Port, Switzerland), SmartSight (Schwind, Kleinostheim, Germany), and Smooth Incision Lenticule Keratomileusis (SILK; Johnson & Johnson Vision, Irvine, CA) (3). These procedures involve the creation and extraction of a refractive corneal lenticule, which is normally disposed of after the procedure (4).

In 2012, we explored the feasibility of cryopreservation of the extracted lenticules for future reimplantation (5, 6). Following successful pre-clinical studies, many authors have demonstrated the therapeutic efficacies of corneal lenticule implantation in patients for the treatment of keratoconus (7–9), hyperopia (10–12), presbyopia (13, 14), and corneal perforations (15, 16). In 2022, collaborating with Cordlife Singapore, a private local lenticule bank, we explored the safety and effectiveness of cryopreserving lenticules in a lenticule bank over 12 months and implanting them into rabbits (17). The study validated lenticule banking service and the product (OptiQ) was subsequently approved by the Ministry of Health of Singapore. In that study, the cryopreserved lenticules were shipped to our institute, stored at 4°C, and implanted within 1 day of delivery. The study showed that although the cryopreserved lenticules had reduced optical transparency when compared to fresh lenticules due to compaction of collagen fibrillar packing, the implant haze largely subsided within 4 weeks after implantation. The interfibrillar distance (IFD) and concomitantly, the transparency were progressively restored to match the recipients’ corneal stroma over 16 weeks. Moreover, there were no adverse reactions or signs of cytotoxicity recorded during the study period.

Here, as a follow-up study, we investigated the ex vivo and in vivo effects of transportation of the lenticules at 4°C in the event of a delay by comparing it to the ideal situation (implantation within a day of shipment). Our rationale was that in smaller countries like Singapore, the transit time of the cryopreserved lenticules is of lesser concern as transportation does not require upwards of 1 day. However, in larger countries like the United States, Australia, India, or China, or if transport is needed internationally, it can take days and can be prolonged by logistic issues or natural disasters. Extended shipping periods can also be expected in places that are difficult to access in vast countries, where infrastructures and supply chain networks are lacking, such as Indonesia or African nations. This knowledge is crucial because effective utilization of the cryopreserved lenticules may be affected by the ability to preserve their structural integrity and transparency over extended transport times. Specifically, we compared ideal (1 day) versus delayed transport periods on lenticular molecular properties and collagen fiber organization and their eventual ramifications on the implantation outcomes in an animal study.

## Methods

### Extraction and collection of fresh lenticules

Fresh lenticules were extracted from 3 donor corneas, procured from Lions World Vision Institute (Tampa, FL). The myopia correction of −6.00D was performed using a VisuMax 500 femtosecond laser system (Carl Zeiss Meditec). The laser parameters included a 100-130 µm cap thickness, a 6.5 mm optical zone with a 6.7 mm treatment zone, and 145 nJ of power with 90° side cut angles. A lamellar dissector (Asico, Westmont, IL) was used to separate the anterior and posterior lamellae of the lenticule, which was then extracted using an EndoGlide microforceps (Network Medical Products, North Yorkshire, UK), through a single incision with 2.1 mm circumferential length, placed at 120°.

### Processing, cryopreservation, and thawing of lenticules

A total of 18 stromal lenticules were donated by nine patients, who underwent SMILE for myopia correction, to Cordlife Singapore. After extraction, the lenticules were placed in their respective sterile containers, filled with saline solution (Opto-Pharm, Singapore), securely closed, sealed in a biohazard bag, and transported at ambient temperature to the Cordlife Singapore facility. Upon arrival at the storage facility, the containers were immediately transferred into a Class II biosafety cabinet that had been disinfected with 70% isopropanol. The lenticules were recovered by pouring their contents into Petri dishes. The lenticules were then disinfected by immersing and rinsing them for 10 minutes in a wash solution composed of Dulbecco’s phosphate-buffered saline (PBS; Biological Industries, Beith HaEmek, Israel) with 1% (v/v) penicillin-streptomycin-amphotericin B solution (Biological Industries). This was followed by a second rinse in fresh wash solution for another 10 minutes. After disinfection, the lenticules were immersed and equilibrated for 10 minutes in a cryopreservation medium composed of Dulbecco’s modified Eagle’s medium, high glucose formulation (Biological Industries), supplemented with 10% (v/v) dimethyl sulfoxide (DMSO; WAK-Chemie, Steinbach, Germany). The lenticules were then transferred to labeled sterile cryovials containing the cryopreservation medium.

All lenticules underwent pre-storage sterility testing by inoculating the cryopreservation medium from the equilibration step into BacT/ALERT BPN and BPA culture bottles, which were then incubated in the BacT/ALERT microbial detection system (BioMérieux, Marcy-l’Étoile, France). Subsequently, the lenticules were subjected to a controlled rate freezing process of −1°C/min until reaching −40°C, followed by a rapid reduction to −90°C at −10°C/min. They were then stored at −196°C in vapor-phase liquid nitrogen tanks for 12 months (n=18). The entire process, from receipt to controlled rate freezing, was completed within 24 to 96 hours.

After 12 months, the lenticules were retrieved from the liquid nitrogen tanks and thawed at 37°C for 1 minute in a water bath. The thawed contents were poured into Petri dishes, and the lenticules were rinsed in freshly prepared wash solution for 10 minutes, followed by a second rinse for another 10 minutes. Post-thaw sterility testing was performed by inoculating the post-thaw cryopreservation medium. The lenticules were then transferred to sterile containers with sterile wash solution, securely closed, and transported on ice to the Singapore Eye Research Institute (SERI) for further experiments. The lenticules were kept in a 4°C fridge at SERI until further use.

### Optical transmittance measurement

Six cryopreserved lenticules were first used to investigate the maximum transportation delay before transparency deterioration: 3 lenticules were stored at constant 4°C to simulate cold-chain management; and the other 3 underwent 40°C incubation for 3 hours daily to simulate potential temperature fluctuations during transportation (Figure 1A). The optical transmittance of the lenticules was measured using an Infinite 200 UV-Vis spectrophotometer (Tecan, Männedorf, Switzerland), with the visible light wavelength ranging from 380 to 780 nm. The lenticules were placed in wells of a 96-well plate (Greiner Bio-One, Kremsmünster, Austria) and inserted into the spectrophotometer chamber for the transmittance measurement. Data were collected at 10 nm wavelength increments. The lenticules were then stored at 4°C until the next transmittance scans or temperature challenge. The scans were conducted at 24-hour intervals. The transparency of fresh lenticules (n=3), serving as the non-cryopreserved control, was also measured.

**Figure 1.**
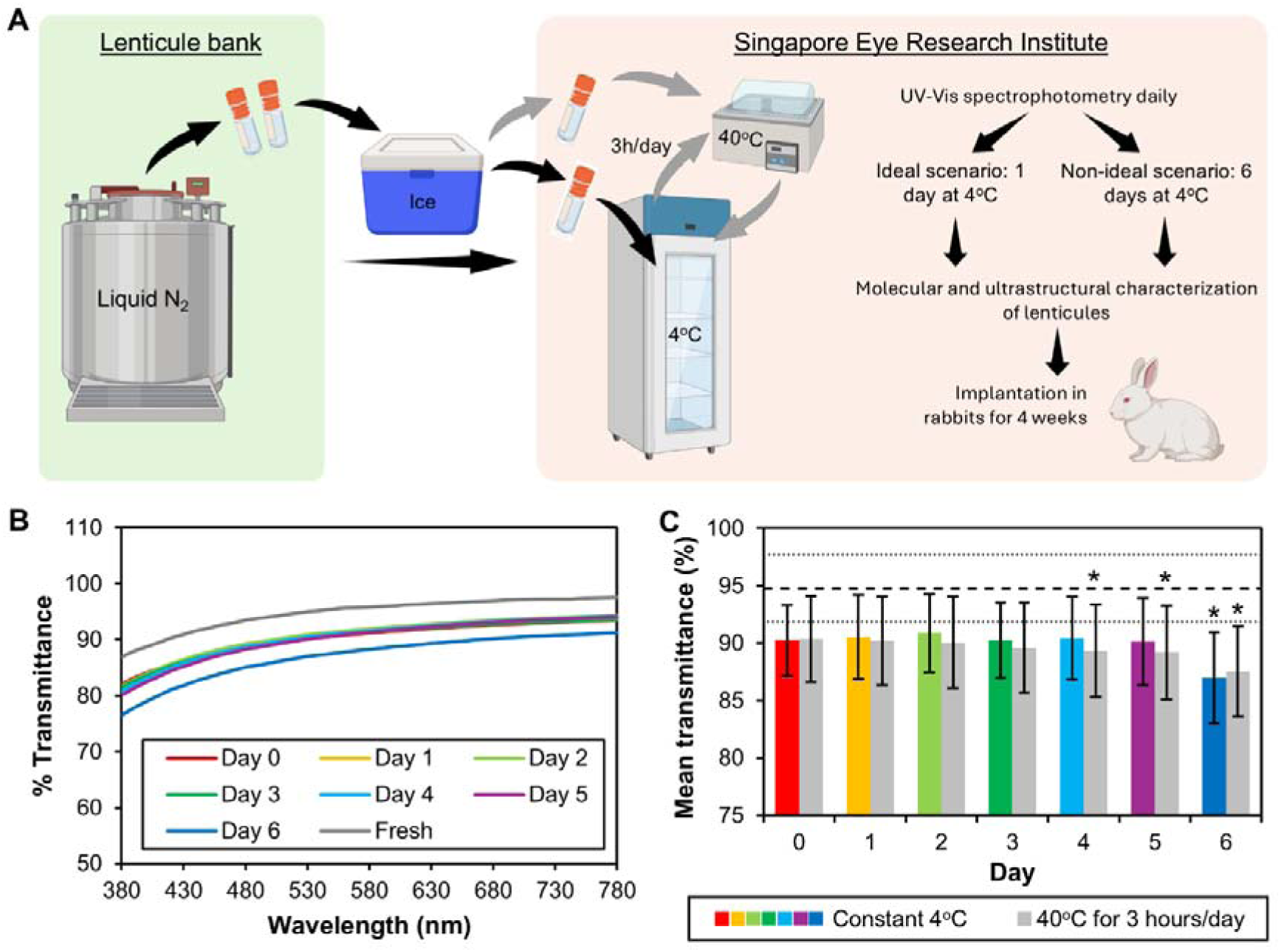
Timepoint determination to simulate ideal and delay in lenticule transportation. (A) Following retrieval from the cryo-storage at the lenticule bank, the lenticules were transported on ice to the lab. The cold chain was maintained then by storing the lenticules at 4°C. Another group of lenticules underwent incubation in a 40°C water bath for 3h daily to simulate potential temperature fluctuations during transportation. The lenticular transparency was measured at arrival (day 0) and daily, up to the point of degradation of the clarity. Ex vivo characterization and in vivo implantation of lenticules, transported following an ideal scenario and non-ideal (transportation delay) scenario, were then carried out. (B) Visible light transmittance scans revealed degradation of light transmittance of lenticules after 6 days. (C) The average transparency up to 5 days was lower than fresh lenticules (the dash line indicates mean and dotted lines indicate standard deviation) but it was not significant. The transparency appeared to worsen substantially on day 6. *p<0.05 vs. fresh.

### Histochemistry

Fresh, day 1, and day 6 lenticules (n=3 in each group) were cut into halves. One-half was subjected to histochemistry and the other to transmission electron microscopy (TEM). Histochemistry of picrosirius red (Abcam, Cambridge, UK), alcian blue (Sigma-Aldrich, St. Louis, MO), and Periodic Acid Schiff (PAS; Abcam) were performed on 8-µm-thick cryosections of the lenticules, according to the manufacturer’s instructions to detect the presence of collagens, proteoglycans, and glycoproteins, respectively. The sections were subsequently viewed with an Eclipse Ti microscope (Nikon, Tokyo, Japan). The intensity of the staining in 3 random fields of view was quantified using ImageJ version 1.54 (National Institute of Health, Bethesda, MD), with results presented in raw integrated density per mm^2^.

### Transmission electron microscopy

The other half of the lenticules and rabbit corneas (n=3 in each group) underwent a sequential fixation process using 2.5% glutaraldehyde (Electron Microscopy Sciences, Hatfield, PA) in 0.1M sodium cacodylate buffer (Electron Microscopy Sciences), followed by 1% osmium tetroxide and potassium ferrocyanide (Electron Microscopy Sciences). The samples were then dehydrated in increasing concentrations of ethanol (Sigma-Aldrich) and two changes of acetone (Sigma-Aldrich), and embedded in Araldite resin (Electron Microscopy Sciences). Ultrathin sections measuring 90–100□nm in thickness were cut using a Leica UC7 ultramicrotome (Leica Microsystems, Wetzlar, Germany). Subsequently, the sections were stained with uranyl acetate (Electron Microscopy Sciences) and lead citrate (Sigma-Aldrich) before imaging with a JEM-2100 microscope (JEOL, Tokyo, Japan). The cross-sectional views of the collagen fibers in the lenticular stroma and rabbit stroma were captured in three random fields. IFD and fibrillar diameter (FD) were then quantified with a minimum of 80 measurements per image using ImageJ version 1.54 (National Institute of Health).

### In vivo lenticule implantation

The in vivo study adhered to protocol 2019/SHS/1470, which received approval from the Institutional Animal Care and Use Committee of SingHealth-Duke-NUS Medical School, Singapore. All animal procedures followed the guidelines outlined in the Association for Research in Vision and Ophthalmology Statement for the Use of Animals in Ophthalmic and Vision Research. Six New Zealand White rabbits aged 9– 12 weeks were sourced from In Vivos, Singapore. Before the experiment, the rabbits were sedated with a combination of ketamine hydrochloride (50□mg/kg; Parnell Laboratories, Alexandria, Australia) and xylazine hydrochloride (5□mg/kg; Troy Laboratories, Glendenning, Australia) via intramuscular injection. Lenticule implantation was performed exclusively in the right eye of each rabbit. Three rabbits received day 1 lenticules, while the other three received day 6 lenticules. The procedure began with the creation of a corneal pocket, followed by incomplete flap creation (flocket) using a VisuMax 500 femtosecond laser system (Carl Zeiss Meditec) (18). The laser setting was: 120□μm flap thickness, 7.9□mm flap diameter, and 170 nJ power. The laser firing sequence was manually halted with 3□s remaining to prevent complete flap creation, leaving only a ∼2□mm circumferential sidecut. Subsequently, a lamellar dissector (Asico) was guided through the sidecut onto the flap bed, releasing lamellar adhesions to create a stromal pocket. Following irrigation with a balanced salt solution, a lenticule was inserted into the center of the flocket, and the opening was closed using interrupted 10–0 nylon sutures (Johnson & Jonhson, New Brunswick, NJ). Sutures were removed after 1 week. The rabbit eyes received Tobradex eye drops (Alcon, Geneva, Switzerland) four times daily until euthanization. The rabbits were euthanized after 4 weeks, and their corneas were harvested and divided equally. One half underwent TEM, while the other half was utilized for immunofluorescence staining.

### Post-surgery corneal imaging

Photographs from a slit lamp, anterior segment optical coherence tomography (AS-OCT), and in vivo confocal microscopy (IVCM) scans were acquired before the lenticule implantation procedure, and 1 and 4 weeks after implantation. Slit lamp images were captured using a Zoom Slit Lamp NS-2D (Righton, Tokyo, Japan). The lenticule’s haze was graded by a masked observer, following our published method (19). Next, corneal cross-sections were captured with AS-OCT imaging, utilizing the RTVue Fourier-Domain OCT system (Visionix, Lombard, IL). The system was adjusted to center the vertex in the AS-OCT image, gradually moved away until the vertical white beam was barely visible, and then the image was captured. Densitometry of the rabbit corneal stroma and implanted lenticule was quantified using ImageJ version 1.54 (National Institute of Health), with results presented in raw integrated density per mm^2^. En-face images of the cornea were obtained using a Heidelberg Retinal Tomograph 3 (HRT3) IVCM (Heidelberg Engineering GmbH, Heidelberg, Germany) with Vidisic carbomer gel (Mann Pharma, Berlin, Germany) as an immersion fluid. The in vivo confocal micrographs were analyzed using Heidelberg Eye Explorer version 1.5.1 software (Heidelberg Engineering GmbH). Visible epithelial wing cells, keratocytes in the anterior half of the stroma, and endothelial cells were enumerated from 3 different areas of the corneas.

### Immunofluorescence staining

Rabbit corneas were harvested and fixed in 4% paraformaldehyde (Sigma-Aldrich) overnight and embedded in an Optimal Cutting Temperature cryocompound (Leica Microsystems). Cryosections of the tissue were cut at 8 um thickness with a Microm HM525 cryostat (Thermo Fisher Scientific, Waltham, MA). Sections on slides were then washed with 1X PBS (1^st^ Base, Singapore) for 5 minutes, thrice and permeabilized with 0.15% Triton X-100 (Sigma-Aldrich) for 30 minutes. Blocking of the unspecific antibody binding was carried out in 2% bovine serum albumin (Sigma-Aldrich) and 5% normal goat serum (Thermo Fisher Scientific) for an hour. Mouse monoclonal primary antibodies (Table 1) were diluted in blocking serum and incubated at 4°C overnight. Slides were washed and a secondary anti-mouse Alexa Fluor 488 antibody (Thermo Fisher Scientific) was then added to incubate for an hour at room temperature. Slides were then mounted with UltraCruz Mounting Medium containing DAPI (Santa Cruz Biotechnology, Dallas, TX). Sections were observed and imaged with a Zeiss AxioImager Z1 fluorescence microscope (Carl Zeiss, Oberkochen, Germany).

**Table 1.** List of primary antibodies.

| Antibody | Clone | Catalog no. | Brand | Dilution |
| --- | --- | --- | --- | --- |
| Fibronectin | FN-15 | F7387 | Sigma-Aldrich | 1:400 |
| Tenascin-c | E-9 | sc-25328 | Santa Cruz Biotechnology | 1:200 |
| Thy-1 | OX7 | sc-53116 | Santa Cruz Biotechnology | 1:50 |
| CD18 | MEM-48 | NB500-379 | Novus Biologicals, Centennial, CO | 1:100 |
| α-SMA* | 1A4 | M0851 | Agilent Technologies, Santa Clara, CA | 1:100 |

\* α-SMA = α-smooth muscle actin

### Statistical analysis

The presentation of data was in the form of mean ± standard deviation. Two-sample Kolmogorov-Smirnov test was carried out to determine the difference in data distribution between the two tested groups. Comparison of multiple experimental groups was performed with one-way ANOVA, followed by post-hoc Tukey tests. A comparison between two experimental groups was conducted with two-tailed independent sample T-tests. Statistical significance was determined at a threshold of p<0.05. The analyses were carried out utilizing Prism GraphPad software version 10.1.2 (GraphPad by Dotmatics, Boston, MA).

## Results

### Transparency and molecular and ultrastructural integrity of lenticules

No microorganism, including fungal, bacterial, or mycobacterial contamination, was encountered in our study (a sample BacT/ALERT test outcome was supplied in Figure S1). Following a year of cryopreservation, on the day of delivery (day 0), the transparency of the lenticules (90.3±3.1%) appeared to be reduced compared to the fresh lenticules (94.8±2.9%), but the decrease was not significant (Figure 1B and 1C). Keeping the lenticules at 4°C for up to 5 days did not cause further deterioration in the transparency (Figure 1C and Table S1). The lenticular transparency became significantly lower than the fresh lenticules after 6 days (87.0±4.0%; p=0.002) (Figure 1C). However, the transparency deterioration was hastened when the lenticules were subjected to a daily higher temperature (Figure 1C and S2 and Table S1). The light transmittance was markedly reduced only after 4 days (89.3±4.0%; p=0.042).

Based on this preliminary transparency test, it was obvious that maintaining the lenticules at 4°C would give us a longer and more acceptable transportation window. Hence, the subsequent experiments were carried out for lenticules transported at constant 4°C. Despite the significant discrepancy in the transparency at 1 day and 6 days, the collagens and glycosaminoglycans (GAGs), which predominantly comprised the extracellular matrix (ECM) of the lenticular stroma (Figure 2A), did not undergo notable changes compared to the fresh and day 1 samples (Figure 2B and Table S2). The glycoprotein content, however, more than the collagens and GAGs, was affected by transportation regardless of the durations. Nonetheless, the difference did not reach statistical significance (Figure 2B and Table S2).

**Figure 2.**
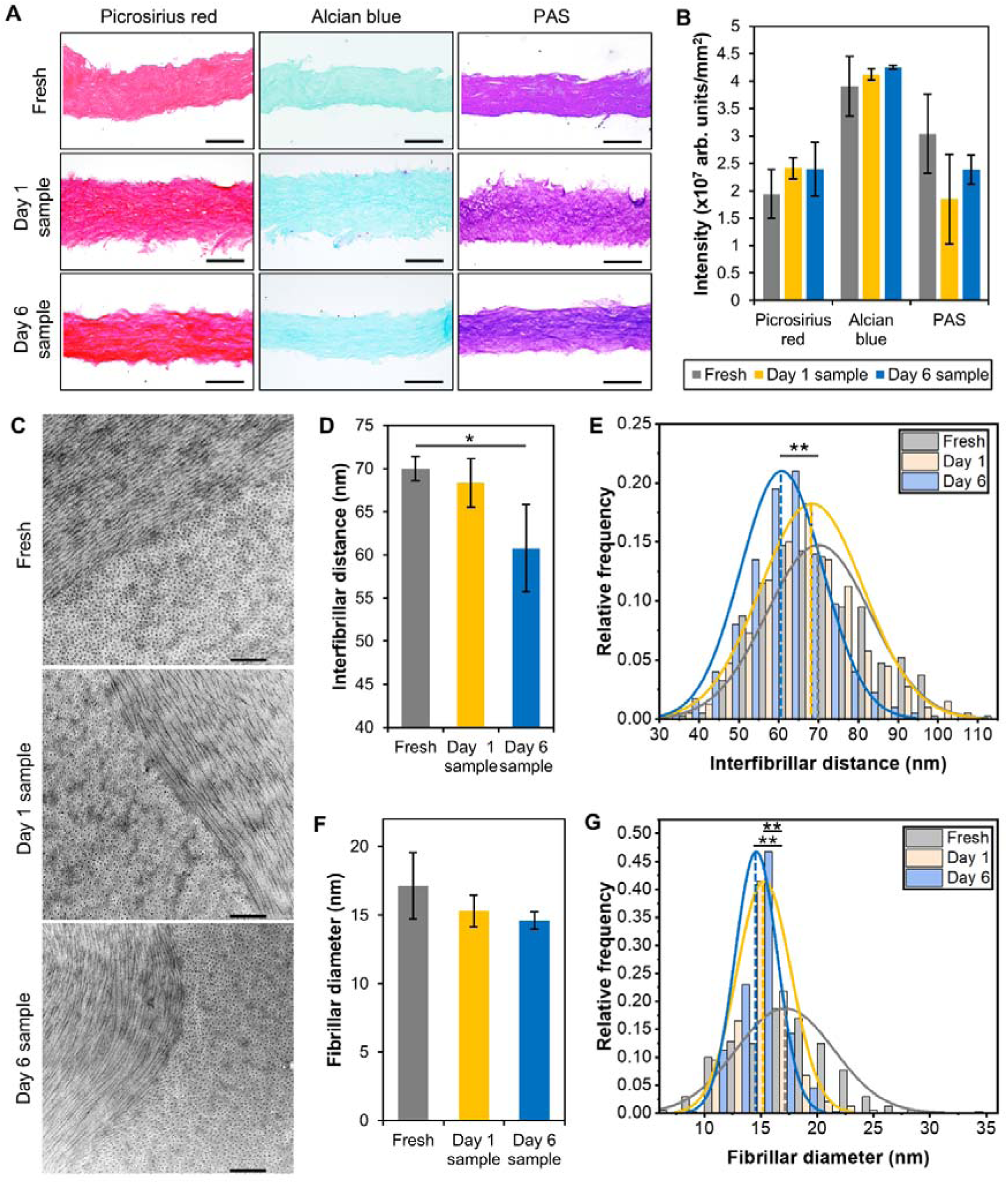
Histochemical and ultrastructural characterization of lenticules. (A) Collagens (stained with picrosirius red), glycosaminoglycans (alcian blue), and glycoproteins (Periodic Acid-Schiff or PAS) were detected in fresh, day 1, and day 6 lenticules. Images shown are in 20x magnification, with scale bars measuring at 50 µm. (B) Quantification of the staining intensity revealed no significant differences between lenticules. (C) Characterization of the lenticules was further carried out with transmission electron microscopy (TEM) to assess the collagen fibrillar arrangement of their stroma Images shown are in 10,000x magnification, with scale bars measuring at 500 nm. (D) On average, the interfibrillar distance was reduced after 1 day in transport, with a further reduction after 6 days. (E) The distribution curve of the interfibrillar distances in the day 1 samples (yellow) shifted from the fresh lenticules (grey) and was increasingly deviated in the day 6 samples (blue). (F) The mean fibrillar diameter was reduced in a similar magnitude in the day 1 and day 6 lenticules. (G) The distribution curves of both groups showed a shift to lower diameters and significantly deviated from the fresh specimens. *p<0.05. *p<0.001.

TEM demonstrated a similar pattern of lamellar stacking in the fresh, day 1, and day 6 lenticules––a lamella comprising longitudinally arranged collagen fibers was followed by cross-sectionally appearing fibers (Figure 2C). Further investigation into the images revealed a 2.3% reduction of the mean IFD of the lenticules after 1 day in transit, compared to the fresh samples (Figure 2D and Table S3). The two specimens appeared to have a similar IFD distribution (Z=1.131; p=0.155) (Figure 2E). The marginal difference in the mean IFD was likely due to the increase in the kurtosis of the distribution (from −0.478 to 0.352), which was contributed by the higher IFD frequency in the 65-70 nm range. The IFD reduction increased to 13.2% and became significant after 6 days in transport (p=0.038) (Figure 2D and Table S3), aligning with the significant transparency deterioration shown in Figure 1A. The IFD distribution pattern was also significantly shifted to lower values (Z=4.419; p<0.001) (Figure 2E). Interestingly, although the FD of day 1 and day 6 lenticules were similarly lowered compared to the fresh samples (Figure 2F and Table S3) and the differences were not statistically significant, the FD distributions were substantially deviated: Z=5.409; p<0.001 for day 1 lenticules and Z=6.435; p<0.001 for day 6 lenticules (Figure 2G).

### Post-implantation evaluations

Slit-lamp photographs of the rabbit corneas, implanted with either day 1 or day 6 lenticules, exhibited no signs of haze and neovascularization in the peripheral regions at any follow-up period, indicating minimal effects on corneal wound healing in the recipients regardless of transportation duration (Figure 3A). The only anomaly observed with the slit lamp was the appearance of haze in or around the implanted lenticules, with day 6 lenticules showing a higher level of light-reflectivity than the day 1 lenticules at postoperative week (POW) 1, where 2/3 of the day 1 lenticules had grade 1 haze and 2/3 of the day 6 lenticules had grade 3 haze (Figure 3B). At POW 4, most of the lenticules in both groups were clear (scored 0), except 1 in the day 6 sample group (Figure 3B). The AS-OCT scans showed that the recipients’ stroma only experienced a marginal but not statistically significant increase in haze at POW 1 (6.9% and 9.5% higher than the preoperative state in the day 1 sample and day 6 sample groups, respectively). The light-reflectivity eventually lowered closer to the preoperative state after 4 weeks (Figure 3C and Table S4).

**Figure 3.**
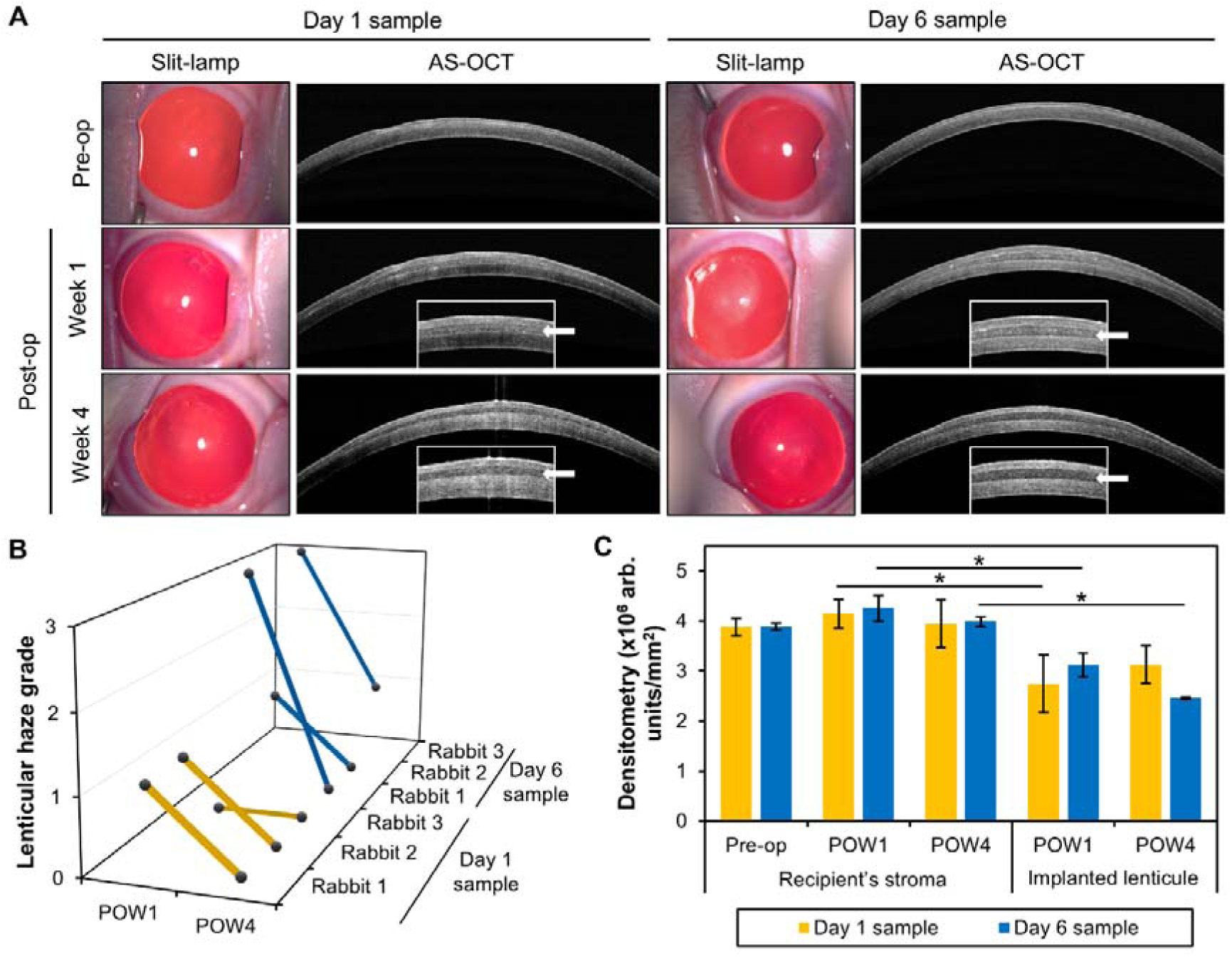
Corneal appearance after lenticule implantation. (A) Slit lamp and anterior segment-optical coherence tomography (AS-OCT) images showed no signs of fibrosis or neovascularization in the rabbit corneas at postoperative week (POW) 1 and POW 4. However, the day 6 lenticules reflected light with greater intensity than the day 1 samples. Insets: Zoomed in AS-OCT images of the central 3 mm of the corneas. Arrows pointed to the implanted lenticules. (B) Grading of the lenticular haze by a masked observer revealed the tendency of a higher score for the day 6 lenticules. (C) At POW 1, the densitometry of the implanted day 1 and day 6 lenticules showed a significantly lower optical density than the surrounding stroma. However, while the density of the day 1 lenticules increased nearer to the stroma at POW 4, the day 6 lenticules became increasingly less reflective. *p<0.05.

Supporting the slit lamp observation above, the AS-OCT-based densitometry showed higher optical density in the day 6 lenticules than in day 1 lenticules at POW 1 (Figure 3A). Nevertheless, the light-reflectivity of both implanted lenticules was still lower than their respective surrounding stromal tissues. At POW 1, the densitometry of both day 1 (p=0.034) and day 6 (p=0.005) lenticules was markedly lower than the recipient’s stroma (Figure 3C and Table S4). Interestingly, at POW 4, the densitometry of the day 1 lenticules increased by approximately 13.9% from POW 1, approaching the optical density of the rabbit stromal tissues (p=0.085). In contrast, the densitometry of the day 6 lenticules continued to decrease and was notably lower than the recipient’s tissues (p=0.001) (Figure 3C and Table S4).

### Tissue responses after lenticule implantation

Despite the differences in the optical density of the lenticules, their implantation did not cause any cellular changes and induce excessive wound healing and inflammation responses in the recipient corneas. At pre-operation, POW 1, and POW 4, en-face IVCM images revealed cobblestone-like epithelial cells, an abundance of keratocytes with elongated nuclei and thin processes in the anterior stroma, and hexagonal endothelial cells in the rabbits (Figure 4A). In addition, the cell number in each layer was maintained throughout the follow-up periods (Figure 4B and Table S5). No significant difference was found in the cell count in any layer compared to the preoperated corneas.

**Figure 4.**
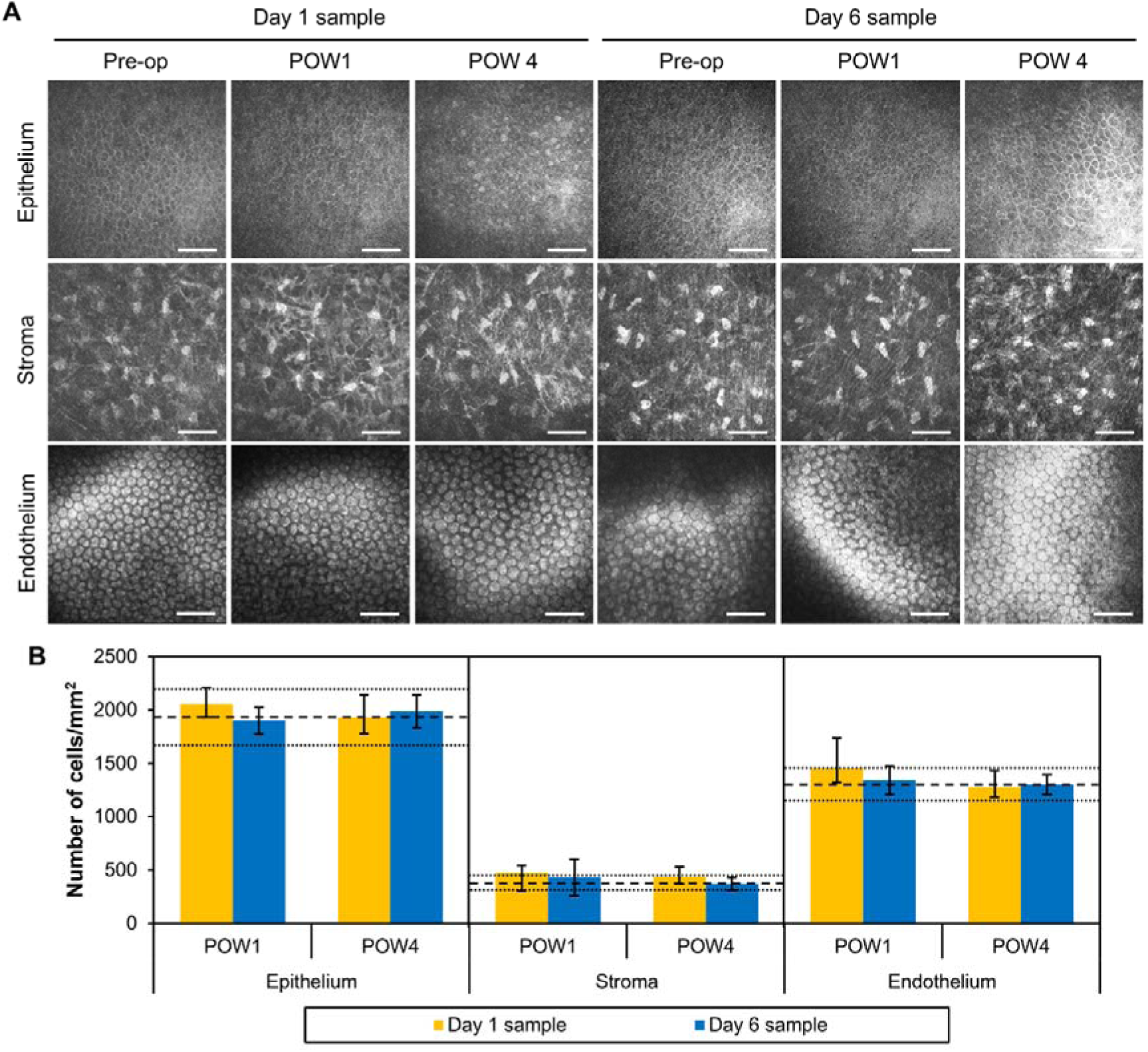
In vivo confocal images of lenticule-implanted corneas. (A) Cells in the epithelial, anterior stromal, and endothelial layers of rabbit corneas implanted with either day 1 or day 6 lenticules showed similar morphology at postoperative week (POW) 1 and POW 4 to that before the implantation. Scale bars = 100 µm. (B) The average cell count in different layers of the corneas was not significantly different from the preoperative corneas (the dash lines indicate means and dotted lines indicate standard deviation).

The immunofluorescence staining complemented the IVCM observation on the minimal tissue reactions caused by the day 1 and day 6 lenticule implantations (Figure 5). Expectedly, the naïve corneas (left-most column in Figure 5) did not express fibronectin, tenascin-C, Thy-1, α-SMA, and CD18. The absence of fibronectin and tenascin across all sample groups suggested that the wound-healing processes were no longer active at POW 4. The absence of Thy-1 and α-SMA, markers for corneal stromal fibroblasts and myofibroblasts (20, 21), respectively, further support the lack of fibrotic responses induced by the implantations. Moreover, CD18 absence indicated no inflammation in the rabbit corneas persisted after 4 weeks. Positive controls were used to demonstrate that the absence of the staining was not due to the inactivity of the antibodies or a staining procedural flaw (right-most column in Figure 5). They were legacy corneal samples harvested from rabbits that underwent irregular phototherapeutic keratectomy or irrPTK after 1 day (22), or chemical burn after 3 weeks (23). Fibronectin, tenascin-C, Thy-1, and CD18 were expressed in the stroma 1 day after irrPTK, while α-SMA was present later after chemical burn.

**Figure 5.**
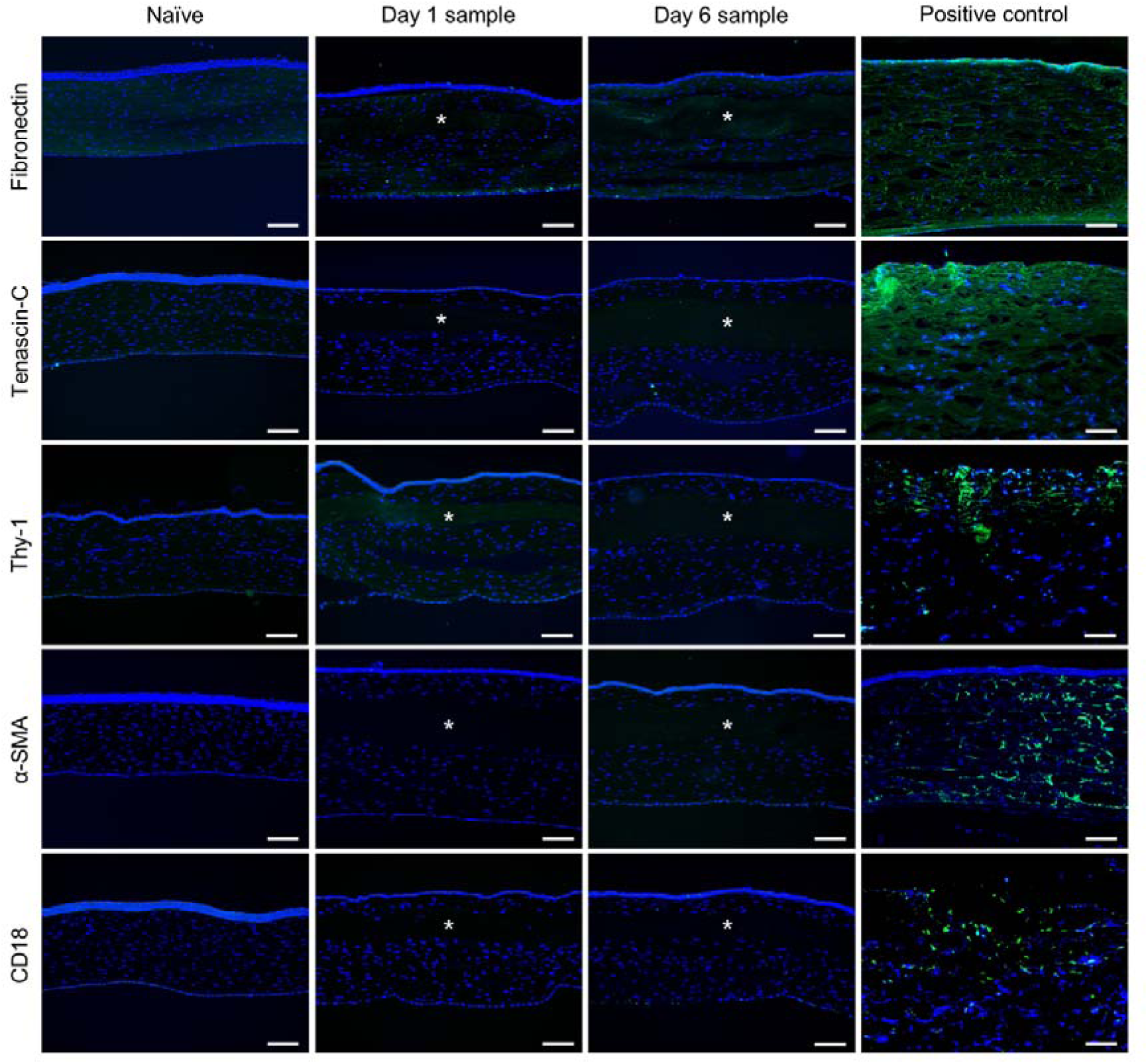
Immunofluorescence staining of lenticule-implanted rabbit corneas. Similar to naïve rabbit corneas, the corneas implanted with day 1 and day 6 lenticules did not induce any fibrosis, marked by the absence of fibronectin, tenascin-C, Thy-1, and α-smooth muscle actin (α-SMA). CD18, a marker of inflammatory cells, was also negative in all samples. Positive controls, namely rabbit corneas a day after irregular phototherapeutic keratectomy after 1 day (for all except α-SMA) or 3 weeks after chemical burn, were used to show that the negative reactivities were not due to inactive antibodies or staining procedural shortcomings. * indicates the lenticules. Scale bars = 50 µm.

### Ultrastructural changes in the stroma after lenticule implantation

Focusing on the ultrastructure of the corneal and lenticular stroma revealed a difference in the collagen fibrillar profile in the implanted day 6 lenticules at POW 4 (Figure 6A). The average IFD (p=0.019) and FD (p=0.004) of these lenticules were significantly lower than the recipient stromal tissues (Figure 6B and Table S6). In contrast, the average IFD and FD of the day 1 lenticules were comparable to the host tissues (Figure 6B and Table S6), with the distribution of the IFD almost overlapping that of the host stroma entirely (Z=1.212; p=0.106) (Figure 6C). The IFD distribution in the day 6 lenticules, on the other hand, was significantly shifted to lower distances (Z=9.144; p<0.001) (Figure 6D). Despite the average FD of the day 1 lenticules not altered, the distribution pattern was still notably different from the surrounding tissues (Z=1.449; p=0.030) (Figure 6E). However, the distribution deviation was not as large and significant as the day 6 lenticules (Z=8.538; p<0.001) (Figure 6F).

**Figure 6.**
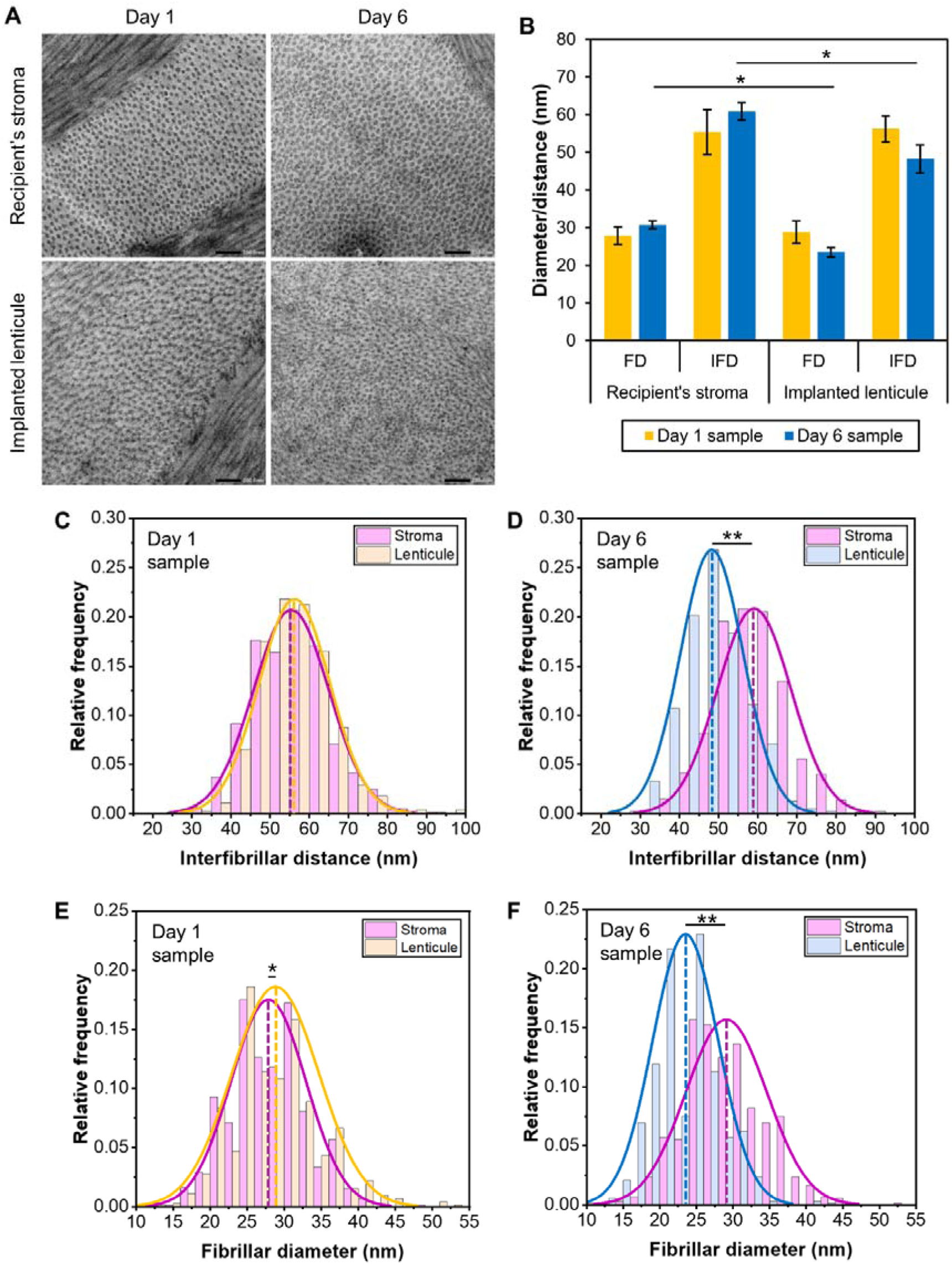
Collagen fibrillar profiles in rabbit stroma and the implanted lenticules. (A) Transmission electron microscopy (TEM) images showed the cross-sections of collagen fibers at 20,000x magnification, with scale bars measuring 200 nm. (B) The average interfibrillar distance (IFD) and fibrillar distance (FD) were similar between day 1 lenticules and the recipient stroma. In contrast, the IFD and FD were significantly lower in the day 6 lenticules than the surrounding tissues. (C) The IFD’s distribution pattern of the implanted day 1 lenticules (orange line) was similar to the host stroma (purple line). (D) In contrast, the distribution was substantially shifted to lower distances in lenticules subjected to delayed transportation (blue line). Although the distribution of the FD in the day 1 samples was altered (E), the deviation was not as large and significant as in the day 6 specimens (F). *p<0.05. **p<0.001.

## Discussion

Our current study provided insights into the long-term cryopreservation (1 year) and clinical viability of cryopreserved KLEx-derived lenticules, especially in scenarios where extended transport times are inevitable. The results of the study suggested that while the ultrastructural and molecular integrity of corneal lenticules could be maintained for short durations (up to five days) in 4°C transportation in PBS, containing penicillin, streptomycin, and amphotericin B, longer transportation times resulted in substantial degradation of transparency and collagen fibrillar ultrastructure. When implanted, the former resulted in earlier lenticular haze clearance and recovery of the fibrillar ultrastructure that closely resembled the recipient’s corneal stroma. In contrast, although the day 6 lenticules became relatively clear after 4 weeks, the fibrillar ultrastructure still significantly deviated from that of the surrounding tissue.

An argument could be made on the superiority of Optisol medium as a storage buffer over PBS for transporting the lenticules (24). After all, donor corneas are transported in Optisol from eye banks (25). Our rationale for the choice was to avoid passing the high cost of Optisol to the patients. Also, because the lenticules are relatively small, they do not require a large volume of Optisol for transportation. Portioning the medium for multiple lenticules is plausible but the quantity of the lenticule retrieval and shipment is currently not large. Reusing the medium later after opening the bottle is not advisable due to contamination risks. Other donor cornea preservatives, such as Life4C and Eusol-C, are similarly costly and share the same aforementioned issues as Optisol (26). Other potential preservatives, although explored as cryopreservation media, such as DMEM, glycerol, silica gel, and silicone oil, could be an option but the outcomes were still inferior to the Optisol (27–29).

Although lenticular transparency was reduced on retrieval after long-term cryopreservation, our findings indicate that an acceptable level of transparency can be preserved for up to 5 days in PBS at 4°C, allowing for feasible transportation in practical settings. After 6 days, a significant decrease in transparency was observed, associated with a marked reduction in IFD and FD. Histochemical analysis revealed that while glycoprotein content was affected, the reduction was not statistically significant and was not unique to the day 6 lenticules; it also impacted the day 1 lenticules. Other molecules, such as collagens and GAGs, were less affected by the cryopreservation and transportation.

We then showed the clinical consequences of implanting these lenticules in the rabbits. These rabbits received topical steroids 4 times a day, similar to the postoperative treatment regime in patients (30). While no neovascularization, overt fibrosis, or cytotoxicity was observed in either group, lenticules that experienced a 6-day delay exhibited a higher propensity for light reflectivity on slit lamp, particularly in the first week after implantation. The lack of vascularization and fibrotic responses in the host corneas, including along the lenticular interfaces, ruled it out as the causative factor for the difference in the lenticular haze resolution. The greater early haze observed in the day 6 lenticules was likely attributed to the greater changes in the IFD and FD than the day 1 lenticules. By POW 4, most of the lenticules in both groups exhibited clear tissues, except 1 lenticule in the day 6 sample group.

It is worth mentioning that the finding on the increased light-reflectivity of the lenticules on slit lamp that did not translate to an increase in the optical density on AS-OCT. In fact, the densitometry of the lenticules in both day 1 and day 6 sample groups showed a decrease compared to the surrounding tissue––a finding consistent with that in humans (7, 9, 12, 30). However, it appeared that while the optical density of the day 1 lenticules was continually elevated nearer to their surrounding tissues, the density of the day 6 lenticules was continually reduced. It is unknown how will this manifest in the patient’s visual quality. Nonetheless, we demonstrated that the discrepancy in the optical density was likely because of the faster collagen fiber organization restoration in day 1 lenticules, which on the TEM revealed that the resemblance of the distribution of the IFD and FD in the day 1 lenticules to that of the surrounding tissues at POW 4––a stromal remodeling phenomenon that we also previously observed after a longer follow-up period (17). In contrast, the distribution of these collagen fibrillar parameters was still notably deviated in the day 6 lenticules compared to the host stroma. Because the animal experiment was limited to 4 weeks, we do not know whether the IFD and FD of the day 6 lenticules would change further due to the continuous stromal remodeling. Considering all the findings suggests that, while cryopreserved lenticules subjected to extended transportation periods may still be usable, the immediate postoperative period may involve greater visual disturbances, which could affect patient satisfaction and visual recovery times.

This study highlights the importance of optimizing the logistics surrounding lenticule banking. The degradation in lenticule transparency after 6 days suggests that lenticule banks should aim to minimize transport delays, particularly for international shipments. Cold-chain management, including strict temperature monitoring, will be critical in ensuring that lenticules arrive at their destination with minimal structural degradation and are implanted within 5 days. The findings from this study are particularly pertinent to the development and operation of lenticule banks in regions where the geographic distribution of patients and clinics necessitates extended transportation times. Future research could explore potential methods, such as the sodium alginate nutrient capsules, containing chondroitin sulfate and sodium hyaluronate (31, 32), to further enhance the stability and transparency of cryopreserved lenticules over extended transport times.

In conclusion, the maintenance of corneal transparency is the barometer for the successful reimplantation of lenticules in vision-restoring surgeries. Corneal clarity is a direct determinant of post-surgical visual acuity, and the observed reduction in optical clarity after 6 days suggests that longer transport times can impact clinical outcomes by delaying visual recovery. The fact that the lenticules maintained an acceptable degree of transparency for up to 5 days in PBS at 4°C after retrieval from the lenticule bank, provides a viable window for lower-cost transportation in real-world settings, provided that proper cold-chain logistics are maintained.

## Data Availability

All data produced in the present study are available upon reasonable request to the authors

## Acknowledgements

The illustrations in Figure 1A were created with BioRender (agreement no. HW26TE6AX5). The authors thanked Mr. Jonathan Miao for his assistance with the imaging analysis. The authors also thanked Cordlife Singapore for the collaboration that entailed the provision and shipping of lenticules.

## Funding resources

The study was supported by the NMRC Clinician Scientist Award-Senior Investigator (MOH-000197-00) and The Lee Foundation Grant (R1917/45/2022).

